# From Delta to Omicron SARS-CoV-2 variant: switch to saliva sampling for higher detection rate

**DOI:** 10.1101/2022.03.17.22272538

**Authors:** Margot Cornette, Bieke Decaesteker, Geert Antoine Martens, Patricia Vandecandelaere, Stijn Jonckheere

**Author notes:** Corresponding author: Stijn Jonckheere, Jan Yperman Hospital, Briekestraat 12, 8900 Ypres, Belgium.

## Abstract

**Background:** Real-time polymerase chain reaction (RT-PCR) testing on a nasopharyngeal swab is the current standard for SARS-CoV-2 virus detection. Since collection of this sample type is experienced uncomfortable by patients, saliva- and oropharyngeal swab collections should be considered as alternative specimens.

**Objectives:** Evaluation of the relative performance of oropharyngeal swab, nasopharyngeal swab and saliva for the RT-PCR based SARS-CoV-2 Delta (B.1.617.2) and Omicron (B.1.1.529) variant detection.

**Study design:** Nasopharyngeal swab, oropharyngeal swab and saliva were collected from 246 adult patients who presented for SARS-CoV-2 testing at the screening centre in Ypres (Belgium). RT-PCR SARS-CoV-2 detection was performed on all three sample types separately. Variant type was determined for each positive patient using whole genome sequencing or Allplex SARS-CoV-2 variants I and II Assay.

**Results and conclusions:** Saliva is superior compared to nasopharyngeal swab for the detection of the Omicron variant. For the detection of the Delta variant, nasopharyngeal swab and saliva can be considered equivalent specimens. Oropharyngeal swab is the least sensitive sample type and shows little added value when collected in addition to a single nasopharyngeal swab.

**Highlights:**

- Saliva is the preferred sample type for Omicron variant (B.1.1.529) detection
- Nasopharyngeal swab and saliva are equivalent for Delta variant (B.1.617.2) detection
- Oropharyngeal swab is the least preferred sample type for SARS-CoV-2 detection

## Background & Objectives

The current standard for SARS-CoV-2 virus detection includes real-time polymerase chain reaction (RT-PCR) testing on a nasopharyngeal swab (NPS), yet the collection of this sample type is experienced uncomfortable by patients. Saliva- and oropharyngeal swab (OPS) collections are considered less invasive. In addition, saliva can be collected by the patients themselves and does not require trained healthcare providers equipped with protective material. OPS is less sensitive as compared to NPS [1], but combining NPS and OPS collection results in a small sensitivity increase compared to the use of a single NPS [1]. Existing literature suggests that saliva can be used as an equivalent alternative specimen for the PCR-based detection of the SARS-CoV-2 virus [2–4]. In fact, preliminary data suggest that saliva may show higher sensitivity compared to NPS regarding the Omicron variant [5]. In this study, we evaluated the relative performance of OPS, NPS and saliva for the Delta (B.1.617.2) and Omicron (B.1.1.529) SARS-CoV-2 variants.

## Study Design

Adult patients who presented at the screening centre in Ypres (Belgium) for a SARS-CoV-2 RT-PCR test between the 3th of December 2021 and 15th of February 2022 were asked to voluntarily participate in the study. Informed consent and questionnaire (assessing eating, drinking, chewing or smoking 30 minutes before test and test indication) were completed under the guidance of the researcher. OPS and NPS were separately collected by the nurse. Based on the Belgian Sciensano guidelines for saliva collection [6], the participant collected the saliva sample in a CE-labeled sterile buffer-free recipient by spitting. No saliva swabbing device was used. Saliva samples were diluted ½ using Sputasol (ThermoFisher Scientific, Waltham, USA). All three samples were analysed separately using the SGRespi™ Pure kit on Maelstrom 9600 (TANBead, Taoyuan, Taiwan) for nucleic acid extraction and Allplex ™ SARS-CoV-2 Assay (Seegene, Seoul, Republic of Korea) on CFX96 Thermocycler (Bio-Rad, Hercules, USA) for PCR-based detection of the SARS-CoV-2 virus. Due to the high specificity of RT-PCR, participants were defined as having a SARS-CoV-2 infection when either saliva, OPS or NPS was positive. To determine the SARS-CoV-2 variant for each participant, SARS-CoV-2 whole genome sequencing (WGS) and clade calling [7] (Ct value < 25) or Allplex SARS-CoV-2 variants I (E484K, HV69/70del, N501Y) and II (K417N, K417T, L452R, W152C) Assay (Seegene, Seoul, Republic of Korea) (Ct value ≥ 25) was performed on the NPS or saliva (sample with lowest Ct value). In brief: WGS was performed using the Research Use Only AmpliSeq for Illumina SARS-CoV-2 Research Panel on Illumina MiSeq (80 samples on V2 flow cell) according to the manufacturer’s standard protocol: 8 µl RNA extract was reverse transcribed using Lunascript RT SuperMix Kit (New England Biolabs, MA, USA), followed by amplification of 237 virus specific amplicons covering > 99% of the 30kb reference genome aiming at a median coverage above 500x, minimal coverage for mutation calling of 10x and variant allele frequency > 90% and a minimum of 30,000 reads per sample and a maximum of 1kb bases below minimal coverage. A consensus sequence was constructed using an in-house pipeline containing Trimomatic for trimming, alignment by BWA, mutation calling by Freebayes and inspection of sequence quality by IGV. Clade calling on the consensus FASTA was done by both Pangolin lineage assignment (v2.3.2) and Nextclade (v0.14.1) webtools. All sequences were uploaded to GISAID.

## Results

We included 246 participants (124 men and 122 women) with median age 39 years (IQR 18.75) of which 155 (63%) participants were SARS-CoV-2 positive on at least one of three sample types. Indications for testing were symptoms (62%) or high risk contact (38%). Comparing the Ct values of the three collected sample types revealed that Ct values are statistically significantly higher for OPS compared to NPS and saliva (figure 1A, ANOVA, p<0.001), reflected by remarkable lower sensitivity and negative prediction value (NPV) regardless the variant type (table 1). Except for three out of 155 infected participants, all patients that tested positive on OPS also tested positive on NPS, resulting in a modest increase in sensitivity when combining OPS/NPS compared to NPS alone (93.5 % vs. 92.2 %, McNemar test, P= 0.5) (table 1). To understand the impact of SARS-CoV-2 variant on performance of sample type, we compared performance of NPS and saliva in both variant groups (Delta, n=62 ; Omicron, n=91; undetermined, n=2; supplementary table 1). Statistically significant lower Ct value for all tested genes was detected in NPS as compared to saliva for both Delta and Omicron variants, with the exception of the E-gene in the Omicron group (figure 1B). However, sensitivity and NPV for Delta variant detection is comparable between NPS (96.8%) and saliva (98.3%). Interestingly, a higher sensitivity is observed using saliva (97.8%) compared to NPS (89.0%) for the detection of the Omicron variant (table 1). Indeed, no statistically significant difference in Delta variant detection between NPS and saliva was observed (McNemar test, p=1), while a statistical significant higher detection rate was observed for the Omicron variant using saliva as compared to NPS (McNemar test, p=0.021). Furthermore, 13 participants tested negative on NPS but positive on saliva (two Delta and ten Omicron). From these 13 participants, a follow-up sample was taken two to four days later, and 11 patients tested positive on follow-up of which 10 were positive on NPS in addition to saliva. Logistic regression showed no impact of eating, drinking, chewing or smoking 30 minutes before the test on SARS-CoV-2 positivity rate for the three sample types.

**Figure 1.**
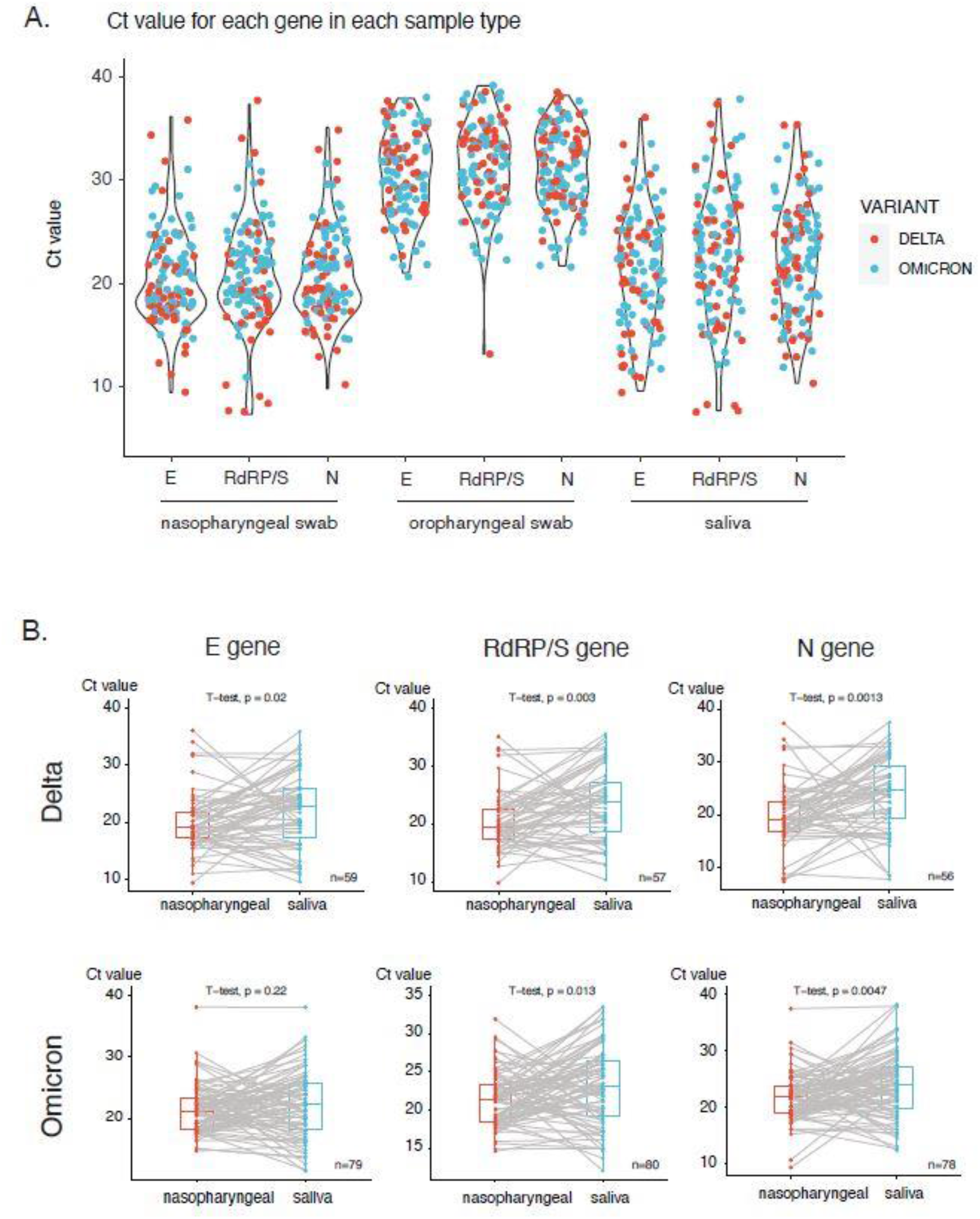
**A**. Ct value for each gene (E, RdRP/S and N-gene) for the three sample types (nasopharyngeal swab, oropharyngeal swab, saliva) (n=114). Thirty-nine samples were omitted due to lack of Ct values for all genes detected in all sample types. Statistically significant difference in Ct values were found for oropharyngeal swabs as compared to nasopharyngeal swabs and saliva for all genes (ANOVA, p<0.001). Statistically significant difference in Ct values were found for nasopharyngeal swabs as compared to saliva for the RdRP/S gene (ANOVA, p<0.001; post-hoc Tukey test, p=0.004) and N gene (ANOVA, p<0.001; post-hoc Tukey test, p=0.014). **B**. Ct value for each gene in nasopharyngeal swab *versus* saliva for SARS-CoV-2 Delta (n=62) and Omicron (n=91) variants. Ct value for nasopharyngeal swab is significantly lower as compared to saliva sample for RdRP/S and N gene in both variants, as well as for the E gene in Delta variant (paired t-test).

**Table 1.** Sensitivity and negative prediction value (NPV) and their 95% confidence interval of nasopharyngeal (NPS), oropharyngeal (OPS), saliva (SAL) sample types or the combination of NPS/OPS for all participants (n=155) and subdivided into Delta (n=62) and Omicron (n=91) SARS-CoV-2 variants.

|  |  | Sensitivity |  |  | Negative Prediction Value |  |
| --- | --- | --- | --- | --- | --- | --- |
|  |  | Sensitivity (%) | 95% CI | number/total | NPV (%) | 95% CI |
| All<br>(n = 155) | NPS | 92,21% | [86,78%-95,90%] | 142/155 | 88,46% | [81,66%-92,96%] |
|  | OPS | 81,17% | [74,09%-87,01%] | 125/155 | 76,03% | [69,56%-81,49%] |
|  | SAL | 98,70% | [95,39%-99,84%] | 152/155 | 97,87% | [92,07%-99,45%] |
|  | NPS/OPS | 93,50% | [88,38%-96,84%] | 144/155 | 91,00% | [83,48%-94,37%] |
| DELTA<br>(n = 62) | NPS | 96,77,% | [88,83%-99,61%] | 60/62 | 96,72% | [88,30%-99,14%] |
|  | OPS | 79,03% | [66,82%-88,34%] | 49/62 | 81,94% | [73,68%-88,04%] |
|  | SAL | 98,34% | [91,34%-99,96%] | 61/62 | 98,33% | [89,41%-99,76%] |
|  | NPS/OPS | 96,77% | [88,82%-99,61%] | 60/62 | 96,72% | [88,30%-99,14%] |
| OMICRON<br>(n = 91) | NPS | 89,01% | [80,71%-94,60%] | 81/91 | 76,19% | [64,07%-85,17%] |
|  | OPS | 83,52% | [74,28%-90,47%] | 76/91 | 68,09% | [57,30%-77,21%] |
|  | SAL | 97,80% | [92,29%-99,73%] | 89/91 | 94,12% | [80,25%-98,44%] |
|  | NPS/OPS | 91,21% | [83,41%-96,13%] | 83/91 | 80,00% | [67,36%-88,58%] |

## Discussion

In this study, we show that saliva is superior compared to NPS for the detection of the Omicron variant, consistent with the preliminary data shown by Marais et al. [5]. Ten of 91 Omicron positive patients tested positive on saliva and negative on NPS, but presented two to four days later with a positive test on both specimen. This suggests that RT-PCR on saliva sample is more sensitive in an early phase of Omicron infection, which might be explained by altered entry pathways and viral shedding of this variant [8–11]. In addition, we show that OPS is the least sensitive sample type for the PCR-based detection of SARS-CoV-2 virus and that combining OPS with NPS has little added value, as was already described by Wang et al. [1]. In summary, our data show that NPS and saliva can be considered equivalent specimen for the detection of the Delta variant, but saliva is the preferred sample type for detection of the Omicron variant.

## Data Availability

All data produced in the present work are contained in the manuscript

## Ethical Approval Statement

The Study Protocol was approved by the Ethics committee of AZ Delta (B1172022000004).

## Author’s Contributions

M. Cornette: conceptualization, methodology, formal analysis, investigation, writing – original draft, writing – review&editing, visualization. B. Decaesteker: methodology, formal analysis, investigation, data curation, writing – review&editing, visualization. G. A. Martens: investigation, resources, data curation, writing – review&editing. P. Vandecandelaere: writing – review&editing, supervision. S. Jonckheere: conceptualization, methodology, writing – review&editing, supervision

