## Supplementary Table 1 for "From Delta to Omicron SARS-CoV-2 variant: switch to saliva sampling for higher detection rate"

Table 1. Distribution of sublineage for all Omicron and Delta variant samples (n=153) determined by WGS (91%), Allplex SARS-CoV-2 variants I and II Assay (5%) or dominant variant at time of sampling (2%). The Allplex SARS-CoV-2 variants I and II Assay is used for determination of variant but not sublineage (undefined).

| **Variant** | **Sublineage** | **Number** | **%** |
| --- | --- | --- | --- |
| **Omicron (n=91)** | BA.1 | 27 | 31% |
|  | BA.1.1 | 34 | 37% |
|  | BA.2 | 24 | 26% |
|  | Undefined | 6 | 7% |
| **Delta (n= 62)** | AY.112 | 2 | 3% |
|  | AY.121 | 2 | 3% |
|  | AY.122 | 2 | 3% |
|  | AY.123 | 3 | 5% |
|  | AY.127 | 5 | 8% |
|  | AY.129 | 3 | 5% |
|  | AY.34 | 1 | 2% |
|  | AY.36 | 3 | 5% |
|  | AY.39 | 1 | 2% |
|  | AY.4 | 3 | 5% |
|  | AY.4.2 | 2 | 3% |
|  | AY.4.9 | 1 | 2% |
|  | AY.43 | 25 | 40% |
|  | AY.92 | 1 | 2% |
|  | AY.98.1 | 1 | 2% |
|  | B.1.67.2 | 1 | 2% |
|  | Undefined | 6 | 10% |
